# A Rule-Based Machine Learning Model for Predicting Virological Failure in Children and Adolescents Living with HIV in Malawi

**DOI:** 10.64898/2026.03.09.26347945

**Authors:** Chimwemwe Chiphe, Thokozani Vallent

## Abstract

Malawi’s HIV treatment monitoring system faces serious challenges because of a shortage of experts and reliance on viral load testing every 3 to 12 months. The process causes dangerous delays in identifying treatment failure. This leads to a higher risk of disease progression, transmission, and death. To tackle this issue, this study used a machine learning model based on association rules and combined it with clustering analysis to create a machine learning framework to identify key factors and risk profiles for virological failure among children living with HIV (CLHIV) in Malawi. The methodology combines a Random Forest classifier for feature importance, association rule mining to find predictive rules, and k-Prototype clustering for risk profiling among CLHIV. The random forest feature importance results show that Body Mass Index (BMI), CD4 count, TB status, ART regimen, gender, ART adherence, and treatment duration are major drivers of virological failure. In addition to these individual factors, the analysis produced highly reliable association rules with over 90% confidence. This establishes a framework for identifying complex risk profiles and informing focused clinical interventions. The high lift values of 4.9 across the most significant rules demonstrate the model’s effectiveness by revealing strong, non-random associations. Clustering analysis also identified two distinct risk profiles associated with virological failure. The k-prototype clustering model performed optimally with a cluster purity of 100% and a silhouette score of 79%.

## 1. Introduction

Human immunodeficiency virus (HIV) is a virus that leads to acquired immunodeficiency syndrome (AIDS), causes failure of the immune system, thus allowing other infections and cancers to affect the body and thrive. The virus attacks the human cluster of differential 4 (CD4 +) cells, causing a decrease in their natural defenses against pathogenic microorganisms (Rosa et al., 2014). The World Health Organization (WHO) recognized that HIV/AIDS-related mortality transmission can be effectively prevented by vigilant monitoring and control of viral load in HIV patients. WHO defines viral load as the quantitative measure of viral particles (copies) per milliliter(mL) in an organism. The results of the HIV viral load testing are categorized as suppressed (less than 1000 copies/ml, indicating no virological failure) or unsuppressed (more than or equal to 1000 copies/mL, indicating virological failure). Elevated viral loads in HIV patients significantly increase the risk of disease transmission and AIDS-related mortality (WHO, 2016).

According to UNAIDS 2022 global HIV statistics, the viral load suppression rate among people living with HIV (PLHIV) was 93%, falling short of the 95% target. While this represents a significant improvement from 83% in 2015, the progress is not consistent across all populations. Among children, for instance, the suppression rate was only 81%, underscoring the difficulty of reaching the global target (UNAIDS, 2024). This disparity highlights the need for intensified efforts to bridge gaps in HIV treatment outcomes worldwide.

Malawi, like other Sub-Saharan African countries, continues to struggle with the HIV pandemic. Although the country is on track to achieve the UNAIDS 95-95-95 goals, virological failure remains a critical challenge in the management of HIV-positive children. CLHIV lags behind adults in viral load suppression (Payne, D et al., 2023). The MPHIA report indicates that Malawi’s overall viral load suppression rate was 94% in 2020-2021. This aggregate figure, however, masks a significant disparity: while adults surpassed the 95% target with a rate of 96.9%, the rate for children lagged at only 81% (MPHIA, 2022). This disparity is particularly concerning, as the viral suppression rate for children has remained below the target since 2015. In fact, the MPHIA 2015-2016 report showed that only 57.9% children achieved viral suppression, compared to 91.3% adults with the target of 90% (MPHIA 2015-2016, 2018). This significant gap in treatment outcomes between children and adults is a concerning trend that needs attention.

As part of routine care in HIV programming in Malawi, healthcare providers routinely assess patients for potential failure of ART treatment. Due to a shortage of experts and the increasing number of cases, manually analyzing patients for treatment failure is impractical (Ndhlovu & Munthali, 2024). Malawi’s current viral load monitoring system for HIV patients has significant limitations. Patients are tested every 3, 6, or 12 months, depending on their viral load status, following the 2016 WHO guidelines. In addition, the results of the viral load test are not provided in real time, leading to long delays and significant changes in a patient’s condition, making timely interventions challenging (Payne et al., 2023).

The lack of predictive models in Malawi’s healthcare system prevents early identification of virological failure, increasing the risk of disease progression, viral transmission, and even death (Ndhlovu & Munthali, 2024). However, machine learning can address this challenge. Machine learning algorithms can analyze complex datasets, identify patterns, and predict future treatment outcomes with high precision (Siddiq, 2022). To address this gap, this study employed an association rule-based machine learning model to uncover complex combinations of factors that predict virological failure. Furthermore, to complement the insights gained from association rule mining, clustering analysis was applied to identify distinct risk profiles among children with virological failure.

## 2. Methods

### 2.1 Ethics Statement

This study received approval from the Malawi University of Science and Technology Research Committee (MUSTREC) under reference number P.03/2025/407. The dataset used for this research came from the Department of HIV and AIDS (DHA), which oversees national HIV data in Malawi. The DHA, along with Global Health Informatics, which manages the database for them, granted access to the data. To protect the privacy and confidentiality of children living with HIV (CLHIV), all direct personal identifiers, including names and ART numbers, were completely removed from the dataset before the research team accessed it. Additionally, the final dataset was secured with a strong password. This way, only authorized researchers with the password could access the data for analysis.

As this research involved a retrospective analysis of fully anonymized program data that was regularly collected and did not involve direct contact with human subjects, the ethics committee waived the requirement for individual informed consent. All methods were carried out according to the relevant guidelines and regulations.

### 2.2 Study Design

The study used a design science research methodology (DSR), which provided the structural framework for the investigation. The process involved repeated cycles of identifying problems and setting objectives, leading to the design and development of an artifact. This artifact was developed to detect actionable patterns related to virological failure (VF) and support its potential prediction among CLHIV. In addition to association rule mining, the study employed clustering techniques, specifically the k-prototypes algorithm, to identify characteristic profiles associated with virological failure among CLHIV.

#### 2.2.1 Association Rule Mining

The apriori algorithm, a part of association rule mining, was used to identify the relationship between selected features and the target feature (virological failure). To generate association rules, the study focused on features related to the target (Antecedent ⟹ Consequent), which helps classify all variables that lead to virological failure (Khare and Gupta, 2016).

#### 2.2.2 K-Prototypes Clustering Method

The study used a clustering method of k-prototypes to identify distinct clinical profiles that require differentiated intervention strategies. K-prototypes combines the k-means and k-modes methods, which are clustering methods for numeric and categorical attributes, respectively (Shahapure & Nicholas, 2020).

### 2.3 Data used for Model Development

The research used viral load data for CLHIV generated between 2022 and 2023. The dataset was obtained from the Department of HIV and AIDS (DHA), the custodian of national HIV data in Malawi. Access to the data was granted by the DHA in conjunction with Global Health Informatics, which manages the database on its behalf.

### 2.4 Data Preprocessing

The pre-processing of the viral load dataset followed a clear pipeline to ensure analytical quality and valid association rules and risk profiles. The process began with data cleaning, where duplicate records and empty rows were systematically removed. This ensured that the dataset contained only complete and unique entries. It helped reduce redundancy and minimize potential biases that could affect itemset frequencies or rule significance.

After data cleaning, the dataset underwent feature extraction. Continuous variables were transformed into categorical attributes through binning. This step allowed clinically relevant patterns to be captured in a way suitable for association rule mining. Specifically, CD4 count was turned into a categorical feature by classifying patients into Low and High immune status. Body Mass Index (BMI) was grouped into ordinal categories: underweight, normal, overweight, and obese. Treatment-related variables were also simplified: ART duration was divided into four intervals (<12 months, 1–3 years, 4–5 years, >5 years), ART adherence was categorized as Poor (<80%) or Good (>80%), and age at visit was divided into developmental groups (<5, 5–9, 10–14, and 15–17 years).

The final step involved feature selection, where only the most relevant attributes were kept for the mining task. This careful selection reduced noise and dimensionality, ensuring that the following rule mining focused on factors with direct significance. The random forest feature importance was used for selecting features.

### 2.5 Machine Learning Model Development Process

#### 2.5.1 Association Rule Mining Development Process

The cleaned data was transformed into a boolean matrix format which is appropriate for computational analysis. The transaction encoder was used whereby, each distinct Feature=Value item was allocated a column. This produced a one-hot-encoded data frame that was suitable for the Apriori algorithm.

The apriori algorithm efficiently identifies frequent itemsets by leveraging the downward closure property (the “Apriori principle”), which states that all subsets of a frequent itemset must also be frequent (Altaf et al., 2017). The algorithm was executed on the encoded transaction matrix with a minimum support threshold of 0.00095. This threshold was selected to identify itemsets that occur with sufficient frequency to be considered nonrandom.

From the generated frequent itemsets, association rules of the form X → Y (where X is the antecedent and Y is the consequent) were derived. Rules were generated based on the confidence metric, with a stringent minimum threshold of 90%. Confidence measures the conditional probability that a transaction containing the antecedent X will also contain the consequent Y. A high confidence threshold ensures that the extracted rules are highly reliable (Agrawal et al., 1996).

The quality of each association rule was evaluated using three key metrics that were automatically generated:

1. Support: The joint probability of both the antecedent and consequent occurring together.
2. Confidence: The reliability of the inference (P(Y—X)).
3. Lift: A measure of the strength of the association, calculated as P(Y—X) / P(Y). A lift value greater than 1 indicates that the antecedent and consequent are positively dependent and the rule is potentially useful for predicting the outcome. A lift value equal to 1 indicates statistical independence between the antecedent and consequent, meaning their co-occurrence is no more frequent than expected by chance. A lift value less than 1 indicates a negative dependence, suggesting the antecedent may be associated with the absence of the consequent (Altaf et al., 2017).

#### 2.5.2 Clustering Model Development Process

To identify distinct CLHIV profiles, the k-prototypes algorithm was employed, as it can handle datasets containing both categorical and numeric variables. The data was converted to a NumPy array for compatibility and the categorical column indices were provided to the algorithm. The optimal number of clusters was determined by iteratively running the k-prototypes algorithm with different values of k. The Cao method or index was employed to optimize the initial centroids for each iteration, efficiently handling the mixed-type data structure.

Once clustering was completed, each CLHIV received a cluster label. Cluster profiles were created to describe the typical characteristics of each group. Numeric features were summarized using median values, while categorical features were summarized using the mode. This profiling method offered a clear overview of each cluster. It highlighted differences in immunological markers, treatment regimens, demographic characteristics, and TB status among patients with virological failure. The performance and quality of the clusters were evaluated using purity and silhouette score.

## 3 Results

### 3.1 Sociodemographic Characteristics of Participants

The study used data from 260 health facilities in eight districts in the southern region of Malawi. From the total population of CLHIV who had ever initiated antiretroviral therapy (ART), a cohort of 5,137 CLHIV currently on ART was analyzed. The data had 56.06% (n=2,880) females and 43.94% (n=2,257) males. Regarding virological outcomes, 81.12% of the cohort achieved virological suppression, while 18.88% experienced virological failure. The 81.12% suppression rate is below the 95% UNAIDS target.

### 3.2 Random Forest Feature Importance Results

The first research objective was to identify the most important predictors of virological failure among CLHIV. A random forest algorithm was used to rank the features according to their importance. Figure 1 gives a summary of the random forest feature importance results.

**Figure 1:**
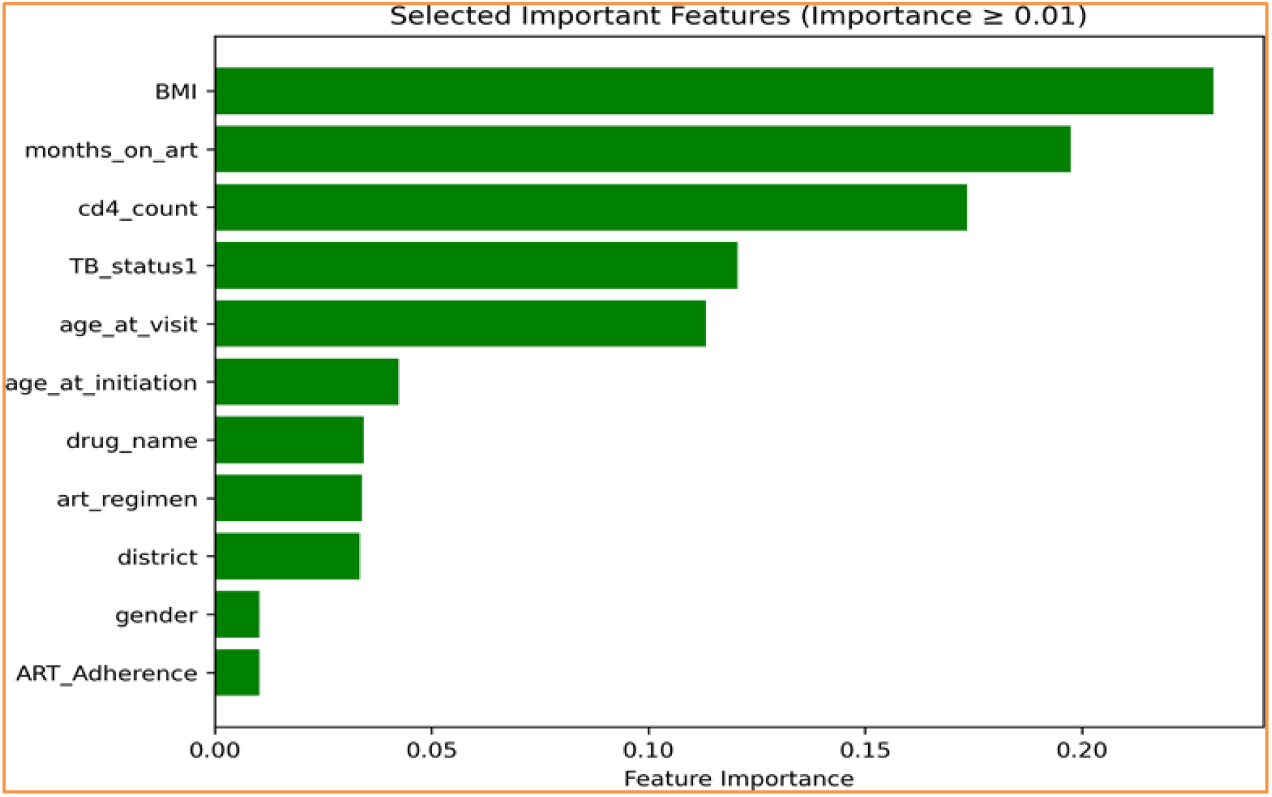
Selected Important Features.

The Body Mass Index (BMI) of CLHIV emerged as the most important predictor of virological failure, with a score of 0.230312. This was closely followed by duration on ART (0.197450). Other significant variables included the CD4 count (0.173426), TB Status (0.120628) and age (0.113225). Variables with lower, but notable, importance scores included age at initiation (0.042483), district (0.046406), ART regimen (0.033972), drug name (0.034407), gender (0.010352) and ART adherence (0.010277).

### 3.3 Association Rule Mining Results and Evaluation

The association rule mining process generated a robust set of 28 rules, which reveal statistical strength as evidenced by key quality metrics. All 28 rules (100%) possess a minimum confidence level of 90%, indicating a very high probability that the consequent is observed when the antecedent is present. This result suggests that the discovered patterns are highly reliable within the dataset. Of all the 28 rules generated, the 10 most important rules were selected to predict virological failure among CLHIV (See figure 2). Below is the presentation of the top 10 association rules generated that highlights interaction of variables that lead to virological failure.

**Figure 2:**
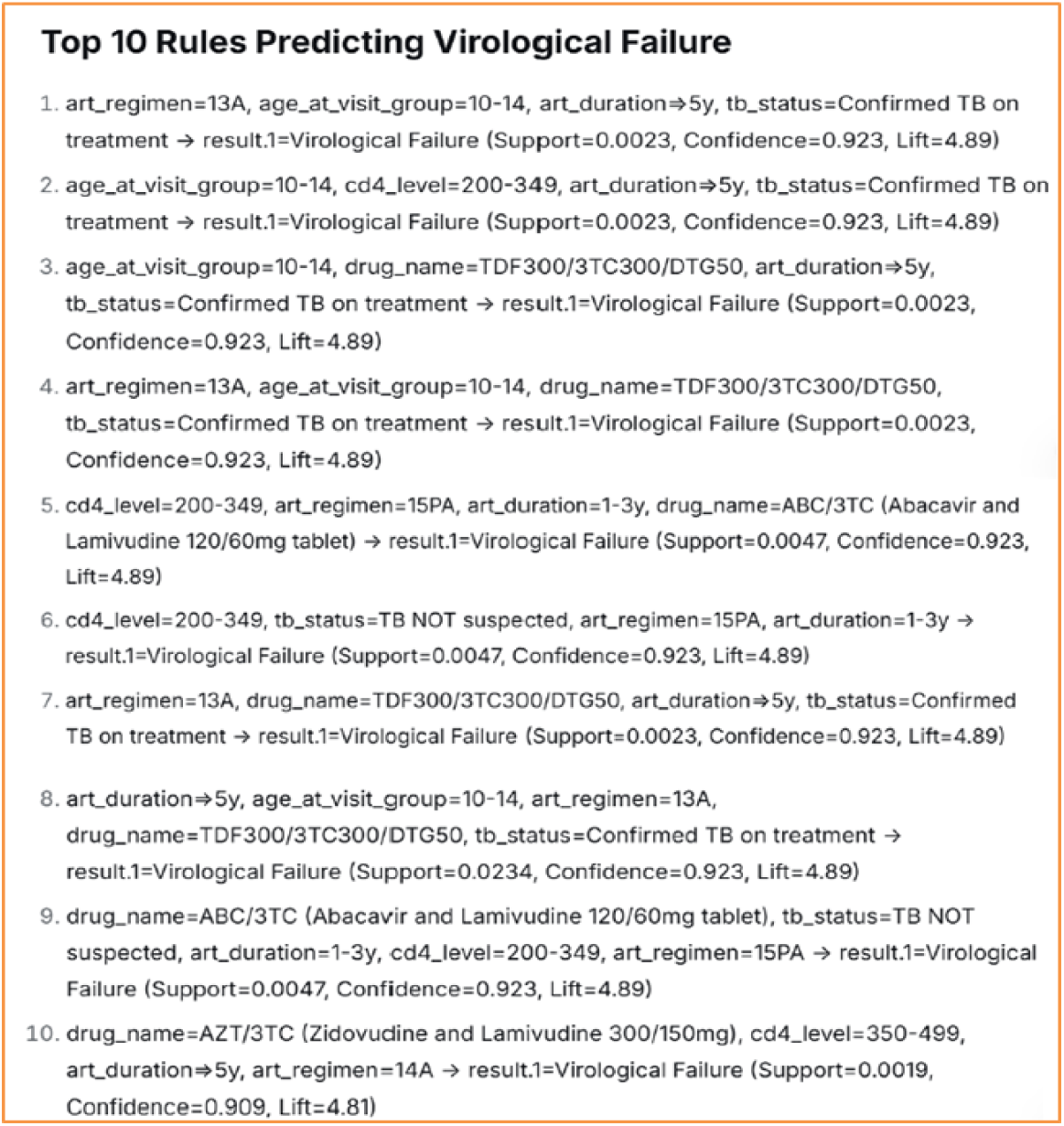
Top 10 Association Rules Predicting Virological Failure Among CLHIV.

**Rule 1** indicated that if CLHIV is on 13A ART regimen, aged 10 to 14, has been on ART for more than 5 years and is confirmed to have TB and on TB treatment. The possibility of developing vilorogical failure is 92.3% of confidence.

**Rule 2** means that if the age of CLHIV is between 10-14, CD4 count is between 200 and 349 cells/mm3, has been on ART for more than 5 years and has confirmed Tb and is on treatment, then the possibility of developing virological failure is 92% of confidence.

**Rule 3** means that if CLHIV with age 10-14 years, using the drug TDF300 / 3TC300 / TTG50, the duration of the ART of more than 5 years, and the TB status is Confirmed TB and on treatment have 92.3% probability of virological failure.

**Rule 4** showed that if a CLHIV is using TDF300 / 3TC300 / TTG50 as drug name, aged between 10-14, TB status is not suspected, has been on ART for period between 1 and 3 years, and the ART regimen is 13A, then the possibility of developing virological failure is 92.3% of confidence.

**Rule 5** means that if CLHIV’ s CD 4 coun t is between 200 and 349 cells/ mm, has been on ART for between 1 - 3 years, is using regimen 15PA with a drug name ABC / 3TC (Abacavir and Lamivudine 120/60mg), then the client will be 92.3% confidence of developing the virological failure.

**Rule 6** indicated that the CD4 level of CLHIV is between 200 and 349 cells/mm3, using ART regimen 15PA, has been on ART for a period of between 1 and 3 years and TB is not suspected, then the possibility of developing virological failure is 92. 3% confidence.

**Rule 7** indicated that if CLHIV is on 13A regimen, the TB status is Confirmed TB and on treatment, using TDF300 / 3TC300 / TTG50 as drug name and has been on ART for period of more than 5 years, then the possibility of developing virological failure is 92.3% of confidence.

**Rule 8** means that CLHIV on 13A regimen, TDF300/3TC300/DTG50 as drug name, has been on ART for over 5 years and TB status is cofirmed TB on treatment, then the CLHIV will have a 92.3% chance of developing the virological failure.

**Rule 9** showed that if a CLHIV’s Cd4 level is between 200 and 349 cells/mm3, has been on ART for a period between 1-3 years, using 15PA ART regimen and TB status is TB not suspected, then the possibility of developing virological failure is 92.3% of confidence.

**Rule 10** means that if a CLHIV’s CD4 level is between 350 and 499 cells/mm3, has been on ART for over 5 years and on 14A ART regimen, then the CLHIV will have a 90.9% chance of developing the virological failure.

The generated association rules demonstrate high predictive reliability, as evaluated by key metrics of confidence, lift, and support. The model was designed to find strong patterns, even rare ones, operating with a minimum support threshold of 0.00095 (0.095%) and a minimum confidence threshold of 90%; this high confidence bar was set based on the established precedent that a rule is considered reliable if its confidence level exceeds 80% (Altaf et al., 2017). With 9 of the 10 main rules boasting a confidence of 92.3%, the model successfully identified specific antecedent conditions that are followed by virological failure with near-deterministic certainty.

**Table 1:**
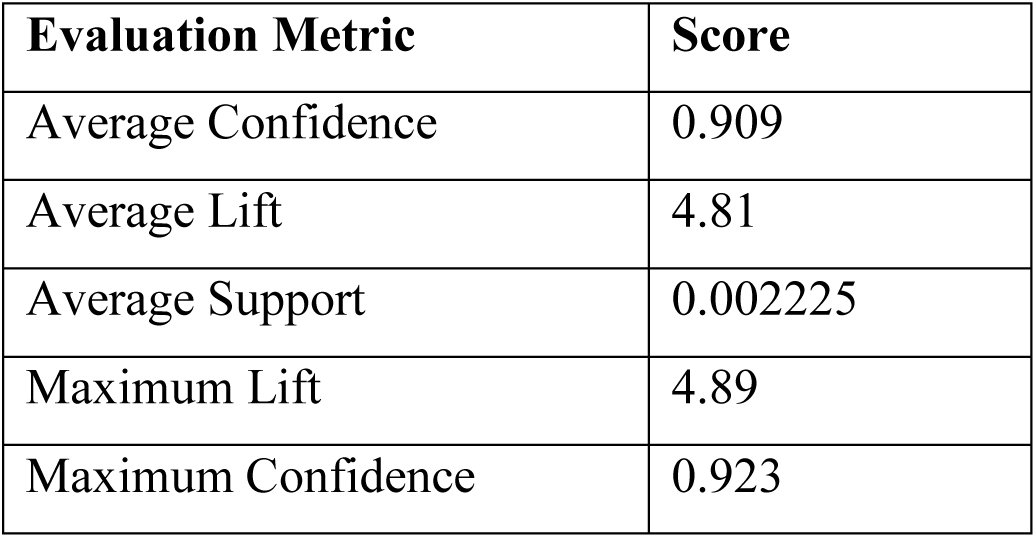
Association Rules Performance.

| **Evaluation Metric** | **Score** |
| --- | --- |
| Average Confidence | 0.909 |
| Average Lift | 4.81 |
| Average Support | 0.002225 |
| Maximum Lift | 4.89 |
| Maximum Confidence | 0.923 |

The average lift of 4.81, with a narrow range peaking at a maximum of 4.89, confirms consistently strong and meaningful associations across all derived rules. The principle of interpreting lift states that a value greater than 1 indicates a positive association between the antecedent and the consequent, while a value below 1 signifies a negative association (Altaf et al., 2017). A lift of 4.9 means that the co-occurrence between the antecedents and virological failure occurs nearly five times more often than would be expected by random chance. See Figure 3 for detailed confidence and lift values for the top 10 rules.

**Figure 3:**
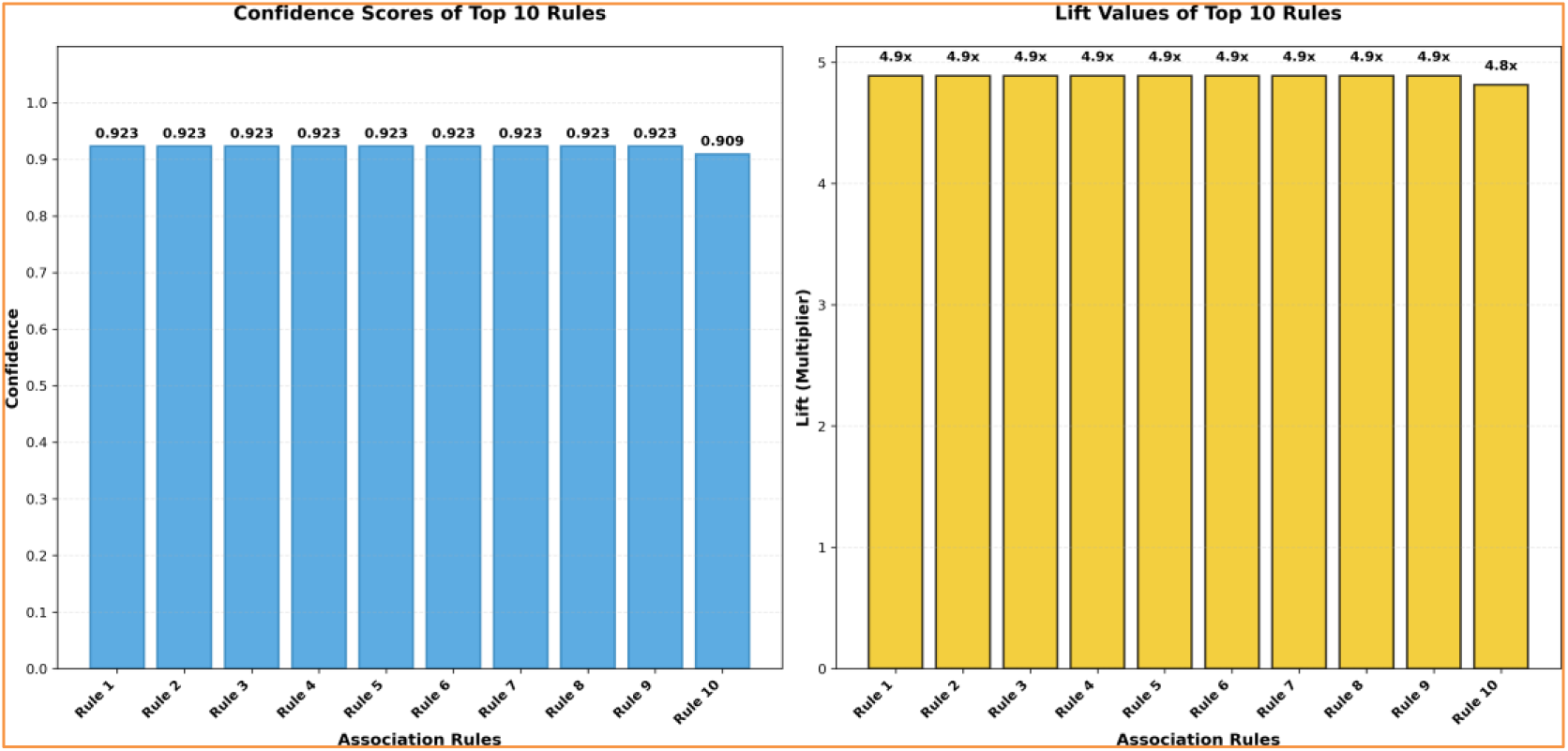
Confidence and Lift values for top 10 rules.

### 3.4 Distinct Risk Profiles for Virological Failure

The study applied k-prototypes clustering to identify distinct risk profiles among CLHIV who experienced virological failure. The analysis generated two clusters, achieving an excellent silhouette score of 79% and 100% purity. This high score signifies strong cluster separation, minimal overlap, and high internal similarity within the identified profiles. In the context of clustering, a silhouette score above 50% is considered acceptable, while a score nearing 70% suggests strong separation, confirming the robustness of the two identified risk profiles.

**Table 2:** K Prototype Cluster Analysis Report.

| Cluster | CD4 Count | Months on ART | BMI | Gender | ART Regimen | Drug Name | TB Status | ART Adherence | Age |
| --- | --- | --- | --- | --- | --- | --- | --- | --- | --- |
| 0 | 1000 | 101 | 15 | Female | 15A | ABC/3TC | TB not suspected | 100 | 12 |
| 1 | 250 | 102 | 16.6 | Female | 13A | TDF300/3TC<br>300/DTG50 | TB not<br>suspected | 100 | 17 |

The distribution of cases across clusters was highly imbalanced, with the 93.7% of observations assigned to Cluster 1. Cluster 0 with 61 observations and Cluster 1 with 909 observations. Both clusters demonstrated 100% purity for the Virological Failure. This indicates that, with respect to the virological result variable, there was no differentiation between the two clusters, all observed cases in the dataset corresponded to virological failure.

The distribution of categorical variables across the two clusters reveals distinct CLHIV profiles. While gender appears relatively balanced in both clusters, suggesting it is not a primary differentiating factor, several other features exhibit pronounced variation. Particularly, tuberculosis (TB) status serves as a strong discriminator, with Cluster 0 comprising exclusively individuals not suspected of TB, whereas Cluster 1, while predominantly consisting of the same group, contains the entirety of the dataset’s TB-suspected and confirmed cases. This clear separation reinforces the clinical relevance of the clustering outcome.

**Figure 4:**
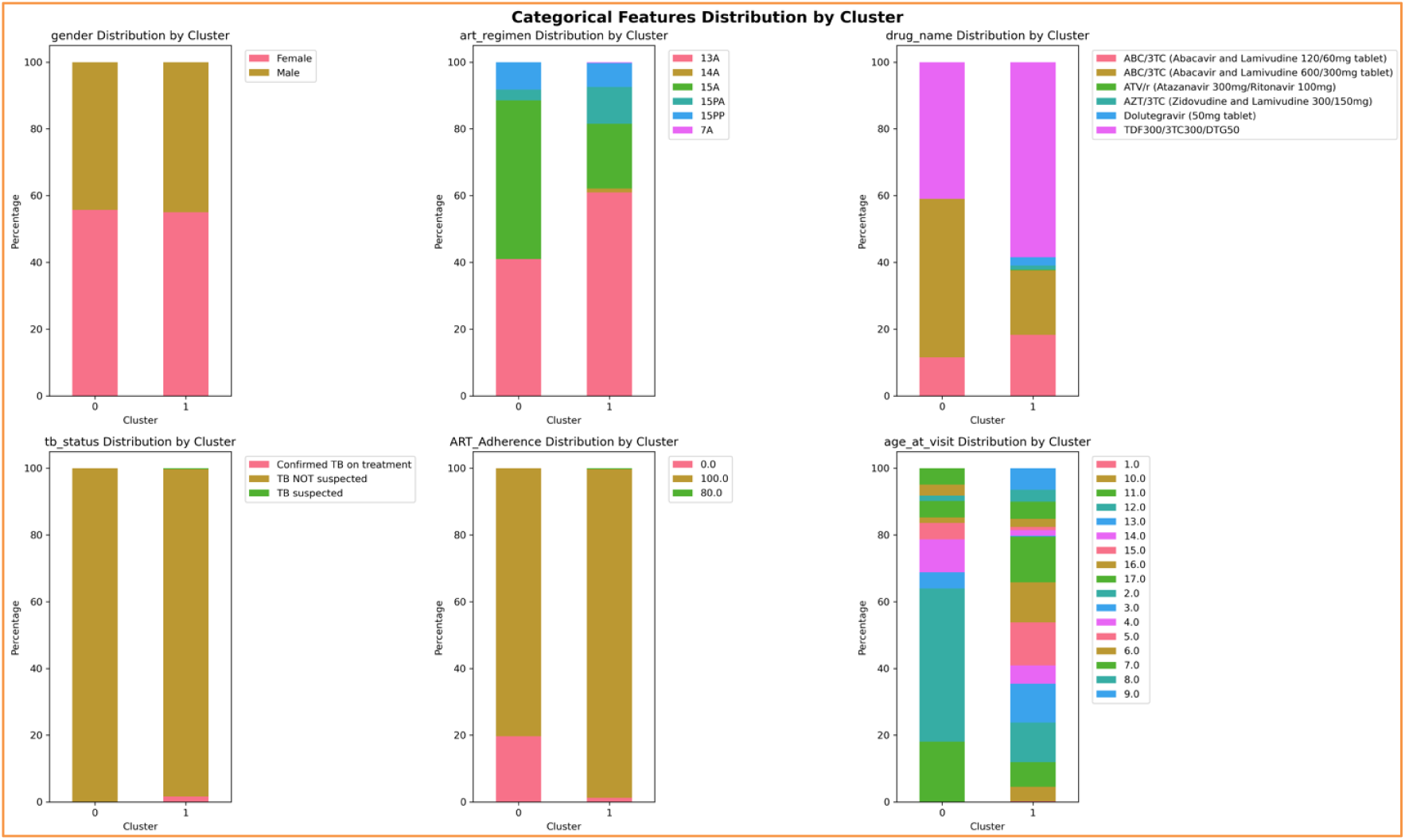
The distribution of categorical variables across the two clusters.

Further differentiation is observed in treatment and geographic patterns. Antiretroviral therapy (ART) regimen distribution varies significantly, with Cluster 0 heavily concentrated on one or two specific drug combinations, such as ABC/TC or AZT/TC-based therapies. In contrast, Cluster 1 demonstrates a broader mix of regimens, including Duloxetine, indicating potential differences in treatment line.

Behavioral and demographic characteristics also contribute to the cluster profiles. ART adherence levels are generally higher in Cluster 0, with a concentration in the upper range (0.8–1.0), whereas Cluster 1 shows a wider and lower distribution of adherence. This pattern may reflect underlying differences in patient engagement, support structures, or clinical management intensity. Additionally, age distributions differ, with Cluster 0 tending toward a younger patient population compared to the broader and older age range present in Cluster 1. Together, these features paint a coherent picture: Cluster 0 represents a more homogeneous, adherent, younger, and geographically concentrated subgroup with no TB suspicion and narrower treatment regimen use. Cluster 1, constituting the majority, is more heterogeneous across all examined dimensions, encapsulating the full spectrum of TB status, regimen type, adherence behavior, age, and region within the cohort.

## 4 Discussion

This study determined the most significant factors that influence virological failure among CLHIV. Among all independent features, BMI, period on ART, CD4 count, current age, age at initiation, district, drug name, ART regimen used, gender, ART adherence, and TB status were the significant predictors of virological failure.

The age of CLHIV is a crucial factor in predicting virological failure. The study found that CLHIV aged 10-14 are at increased risk of virological failure. This is because they are transitioning to independent HIV management and struggle with daily adherence to oral medication, leading to sustained viremia, HIV-related mortality, and transmission to partners and children (Rakhmanina et al., 2024). This finding aligns with studies conducted in Mozambique (Ruperez, 2015) and in Ethiopian localities like Gondar (Bayu et al., 2017; Meshesha et al., 2020), which reported that virological failure occurs more frequently among adolescents living with HIV than among older HIV patients. The likely reason could be the emotional instability of young HIV patients, which can contribute to depression and anxiety about sharing their status with family and friends, negatively impacting treatment outcomes (Meshesha et al., 2020; Fentaw et al., 2020).

Duration on ART was an important feature for predicting virological failure. CLHIV who had a longer duration of stay on ART had a higher risk of developing virological failure. Similarly, previous studies conducted in Waghimra, Northern Ethiopia, and Mettu, South West Ethiopia (Zenu et al., 2021; Emagnu et al., 2020) revealed that patients who had been on ART for a longer time were more susceptible to developing virological failure. This is due to the prolonged use of ART, which increase the risk of developing poor adherence to treatment and unfavorable medication reactions (Ansah et al., 2021).

The CD4 count of CLHIV was another relevant feature for predicting virological failure. As a result, CLHIV with low CD4 counts were more likely to develop virological failure. This finding is supported by research conducted in the Waghimra zone, Northern Ethiopia (Emagnu et al., 2020) and Northeast Ethiopia (Meshesha et al., 2020), which reported that patients with low CD4 counts were more likely to experience virological failure. The effect of low CD4 counts makes patients more susceptible to opportunistic infections, which could increase the chance of virological failure (Ethiopian Federal Ministry of Health, 2021).

This research reveals that CLHIV who have TB are more likely to develop virological failure. This finding is consistent with a substantial body of evidence highlighting the impact of TB co-infection on HIV treatment outcomes. TB co-infection induces a state of chronic immune activation, which may facilitate HIV replication and increase the risk of virological failuire, even when adherence is maintained (Ansah et al., 2021). Studies in sub-Saharan Africa have corroborated this link; for instance, a cohort study in South Africa reported a nearly two-fold increased risk of virological failure in HIV-TB co-infected patients compared to those with HIV alone (Maskew et al., 2022).

The ART regimen was highly important in predicting virological failure. This research showed that patients who were taking the first-line ART regimen of 13A and drug name TDF300/3TC300/DTG50 were more likely to develop virological failure. Findings from a research study which was conducted in Nepal (Chet Raj Ojha et al., 2016) and Namibia (Mouton et al., 2017) supported that patients who received the first-line ART regimen of TDF-3TC-EFV were more prone to virological failure. This idea is also supported by the WHO, which recommends starting ART treatment with DTG rather than Efavirenz (EFV) due to difficulties with the high prevalence of depression, dizziness, and treatment-related adverse effects (World Health Organization et al., 2021).

The gender of CLHIV is another significant factor in predicting virological failure. This study is in line with the research studies conducted in Kumasi, Ghana (Ansah, et al., 2021), Morocco (Hicham, et al., 2019), and the Tigray region, Ethiopia (Desta, et al., 2020) that stated that virological failure was more common in male HIV patients than female HIV patients. The possible reasons might be excessive alcohol consumption, low clinic attendance for ART, and low health-seeking behavior in male patients (Desta, et al., 2020).

Building on the interaction of factors influencing virological failure among CLHIV, our cluster analysis revealed two distinct profiles. Cluster 0 represents younger children (around 12 years old) who are severely underweight and on the 13A regimen, with surprisingly high CD4 counts despite being classified as having virological failure. On the other hand, Cluster 1 represents older adolescents (around 17 years old) with lower CD4 counts who are on the first-line TDF/3TC/DTG 15A regimen. These two clusters suggest two distinct virological failure risk profiles: (1) young, underweight children with unusual high CD4 counts yet experiencing virological failure, and (2) adolescents/older children with low immunity and virological failure.

Age of the CLHIV emerged as the primary driver distinguishing the two CLHIV profiles, with clear demographic and clinical separations between younger children and older adolescents. The analysis reveals that nutritional status, measured by BMI, differs strongly between clusters, while CD4 count provides a sharp biological separation between the groups. Furthermore, ART regimens systematically differ by age group, reflecting standard treatment protocols. The analysis further indicates that Tuberculosis (TB) co-infection is not a significant factor in this cohort’s virological failure. TB cases were found exclusively in Cluster 1, constituting a mere 2% of the cases within that cluster, which underscores its limited role in defining these distinct patient profiles.

### 4.1 Comparative Analysis of Association Rule Mining and Clustering Results

The association rules and clustering results show both similarities and differences regarding virological failure among CLHIV. Rules 1, 2, 3, 7, and 8 align with Cluster 1 of older adolescents (median age 17) on TDF/3TC/DTG regimens. However, there is a notable difference concerning tuberculosis. While the rules consistently recognize confirmed TB co-infection as a strong risk factor, clustering indicates that TB cases make up only 2% of Cluster 1. This suggests that the association rules highlight a specific high-risk subgroup within this broader adolescent profile rather than representing its average characteristics.

A clearer difference appears in the pediatric profile. Rules 5, 6, and 9 focus on younger children on ABC/3TC regimens with moderate immune suppression (CD4 200–349) and no TB suspicion. This partially matches Cluster 0 of younger demographic and regimen characteristics but contrasts with Cluster 0’s unexpectedly high CD4 counts (median 1000). This immunological difference suggests that clustering may have recognized a unique situation of virological failure occurring despite normal CD4 counts, which is not captured by the association rules. Meanwhile, the rules reflect a more clinically typical pattern of failure linked with moderate immunodeficiency.

These findings show that clustering offers a broad classification based on age as the main factor. In contrast, association rules are better at identifying specific, complex risk patterns that may point to important subgroups within or across these larger clusters. This highlights the value of using both methods to understand the variety in virological failure.

## 5 Limitation of the Study

This analysis was limited to clinical data from health facilities and did not account for important social determinants of virological failure in CLHIV, such as patient and caregiver education level, household food security, and economic stability. This is because the health facilities do not collect data on social factors, they only focus on clinical data. Future studies should incorporate both clinical and social factors to better predict virological failure.

## 6 Conclusion

In conclusion, the study demonstrates the feasibility of using machine learning methods to identify CLHIV at high risk of virological failure and to determine predictive factors associated with virological failure. Machine learning algorithms seem to work effectively for risk prediction and classification of ART treatment failure, but further refinements are needed. Nevertheless, the study model may contribute to the important public health issue of reaching and treating CLHIV.

Machine learning can be of great help to clinicians who care for HIV patients. The proposed algorithm can predict viral failure in HIV patients with optimal confidence, support and lift. This prediction helps to optimize the use of hospital resources to treat high-risk patients, provide better quality care, and reduce medical errors caused by fatigue and long work hours in ART clinics. Designing effective predictive models may improve the quality of care and improve patient survival. Therefore, our study of examining interaction of factors that influence virological failure can make a significant contribution to identifying HIV patients at high risk of viral failure and adopting the most effective supportive and therapeutic regimens. This may reduce ambiguity by providing quantitative, objective, and evidence-based models for risk stratification, prediction, and ultimately care planning.

## Data Availability

The data is readily available. The data was collected from the Malawi's department of HIV and AIDs

## Notes

### Competing Interest Statement

The authors have declared no competing interest.

### Funding Statement

The author(s) received no specific funding for this work.

### Author Declarations

Ethics application reference number is P.03/2025/407. MUSTREC reviewed and approved the application through an expedited review process.

